# A Method to Model Outbreaks of New Infectious Diseases with Pandemic Potential such as COVID-19

**DOI:** 10.1101/2020.03.11.20034512

**Authors:** Willem G. Odendaal

## Abstract

The emergence of the novel coronavirus (a.k.a. COVID-19, SARS-CoV-2) out of Wuhan, Hubei Province, China caught the world by surprise. As the outbreak began to spread outside of China, too little was known about the virus to model its transmission with any acceptable accuracy. World governments responded to rampant misinformation about the virus leading to collateral disasters, such as plunging financial markets, that could have been avoided if better models of the outbreak had been available. This is an engineering approach to model the spread of a new infectious disease from sparse data when little is known about the infectious agent itself. The paper is not so much about the model itself - because there are many good scientific approaches to model an epidemic - as it is about crunching numbers when there are barely any numbers to crunch. The coronavirus outbreak in USA is used to illustrate the implementation of this modeling approach. A Monte Carlo approach is implemented by using *incubation period* and *testing efficiency* as variables. Among others it is demonstrated that imposing early travel restrictions from infected countries slowed down the outbreak in the USA by about 26 days.

## I. Introduction

**E**ven before the coronavirus was officially declared a *pandemic*, it was being heavily politicized in the USA already in the midst of a presidential election year. Coloring the outbreak with political slants in the media exacerbated the transmission of misinformation faster than the virus itself. This lead to extreme over- and under reactions in society leading to collatoral calamities including: a stock market crash, unnecessary quarantine measures often involving harsh conditions, a blow to the travel industries, shortages and price gouging of medical supplies, and unnessecary panic in general born out of uncertain expectations.

Health professionals and organizations specializing in epidemics and infectious disease, were hard at work in laboratories focused on researching and gathering information about this new strain of which very little was known, and working on tests, vaccines, and cures. (Fortunately, the People’s Republic of China had shared their sequencing so that other countries could also develop tests for detecting infections.) However, data that are crucial for planning a strategic response to a pandemic at the government level were missing, and remained so for months into the outbreak. Missing data included, latent period, incubation period, level of asymptomatic contagiousness, average time since exposure before it becomes contagious, means of transmission, level of contagiousness, average hospitalization rate, infection rate, fatality rate, and the number of people already infected, the resilience of the virus to a variety of external factors, among others. This is a long list of unknowns and it is incomplete. Many of the variables have to be expanded demographcally. For decisions at the government level in response to the pandemic, good models are necessary from which decent projections can be made.

If better modeling tools had been utilized and prediction widely communicated to governments, the media, and social media well in advance, the severe ramifications could have been avoided, or at least lessened. One of the most sought after estimates is the projected number of people who may are infected on a given day. Many of the infected still fall within the incubation period and may be spreading the virus unknowningly. This parameter is useful for estimating the extent of outbreak among a population over time and what to expect in numbers that follow, such as hospitalizations and fatalities. Models that are suitable for projected estimates are important for common sense decision making by governments, organizations, and even individuals to contain, prepare, prevent, react, and safeguard.

However, good models cannot be developed without good data. And on data collection the system failed miserably. Data from China was widely considered unreliable, except by a few such as the World Health Organization. But at the national level there should have been more proactive steps taken to eliminate the unknowns and collect data. For example, no testing was performed on even one representative population for extrapolation to estimate the infections in the USA. There were no initiatives to conduct testing and lab experiments to learn what the incubation period is, or the resilience of the virus to temperature or humidity, or anything that would have improved the models. Instead, the experts were collecting anecdotal evidence from around the world and making decisions based on the conclusions of non-peer-reviewed articles about the pandemic in Wuhan sources that were known to be unreliable. Without good data there can be no confidence in models from it.

The challenge being addressed herein, is to model the early spread of a new infectious disease knowing very little, other than sparse incoming world data.

## II. Core Ingredients of the Model

This model is characterized by the following

- Purpose
- Assumptions
- Mathematical approximation
- Presentation

### A. Purpose

The main purpose of the model is to predict the spread of a new infectious disease during the very early stages of the outbreak knowing very little, other than sparse collected data such as incoming world data of confirmed infections and confirmed fatalities. Likely scenarios can then be presented based on known and unknown parameters that are clearly defined for societal decision making. Purpose is closely tied to presentation. One of the most important figures for which estimates can be projected using this model includes the unknown number of people who have already been infected on any given day in the future, especially if asymptomatic infections are also contagious during the incubation period, the average length of which may also be unknown.

### B. Assumptions

It is very important to identify, define, and declare all conditions and assumptions in a model. For coronavirus, very little was known, so the following assumptions were made:

#### 1) Unknowns

It is assumed that nearly nothing is known about the disease, except that it is an infectious disease that may or may not have similar characteristics to other known viruses. Unknown characteristics, parameters, variables, statistics, etc. may include among others:

- average incubation period, latent period, etc.
- means of transmission
- the level of asymptomatic transmission, if at all
- resilience to temperature, humidity, etc.
- survival rate on objects and surfaces
- testing accuracy (The percentage of infected people tested, who tested positive.)
- testing efficiency (The percentage of all infected people, who tested positive.)
- reliability of sample data

#### 2) Reactionary Fine-Tuning

As with any model, it should be fine-tuned as more information is gathered. It should also be adjustable to external events that will influence the rate of the outbreak, or dramatic changes in the incoming statistical data.

#### 3) The Law of Averages

For illustration purposes the law of averages will be used. The method can then easily be extended by implementing statistical theory with probabilities and uncertainties to perform regression analysis. However, a statistical approach is essentially pointless given the large uncertainties due to lack of information,

#### 4) Valid Data Groups from global Data Population

For coronavirus the data population consists of case statistics from all over the world. However, when plotting the cumulative number of confirmed cases on the planet as a function of time, the curve is artificially erratic and dominated by unreliable data out of China. To establish a more reliable trend, the data from the PRC was therefore subtracted from the data collected from the rest of the world. It is then assumed that the rate at which the outbreak spreads in most countries should, on average, roughly follow the average rate in the world excluding China.

#### 5) Mathematical Form

Any appropriate mathematical function can be used. This peper is not about the model, but about the approach. To best illustrate the method, the cumulative rate of infections is assumed to be a simple exponential of the form:

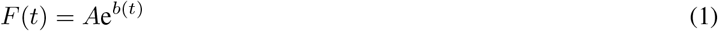

where *A* and *b* are variables to be determined and *t* is time. The number *e* is the base of the natural logarithm equal to 2.71828. This first-order equation is not the only mathematical formula that can be used. It is an approximation which would be limited to the very early stages of an outbreak and is not appropriate for modeling the entire outbreak. A better function for modeling a complete outbreak would be piece-wise function, the logistic function, or one based on the Heaviside equation to incorporate saturation and declining infections.

However, equation (1) has the unique property that it’s time derivative and time integral results in the same function, except scaled by an amount. In other words, the virus is spreading at a rate that accelerates, that the accelerated rate itself accelerates, and so on. It appears as a straight line on a logarithmic scale. Another unique property of this function is that neither linear scaling of it, nor a constant shift in time, changes the time dependent exponent. Thus, time delays and taking percentages are interchangeable and can be transformed one into the other. In fact, derivates, integrals, time shifts, and linear scalling all result in a function of the same form and changes only the coefficient, *A*. Graphically, this means that none of these operations change the slope of the trend line on a logarithmic scale.

Therefore, because of all these handy features, the exponential function will be used for simplicity to illustrate how the model works.

### C. Discretizing the Model

Since data and graphs are best collected and presented by date, time will be quantized as days in *i*, dates by *d* =*day number*, and *time periods* such as time delays also in days, by *p* =*period in days*. The difference between these two is that *d* might, for example, represent a date in the future, while *p* might represent an intrinsic parameter such as the incubation period, or the number of days that the average world spread lagged or lead the first introduction of the virus in a specific country. The function is then rewritten as follows:

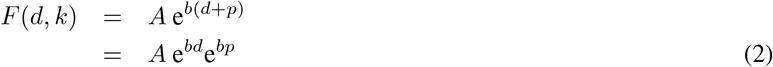

If *F* (*d, k*) represents the number of new cases on a particular day, then its time integral gives the cumulative cases over a period:

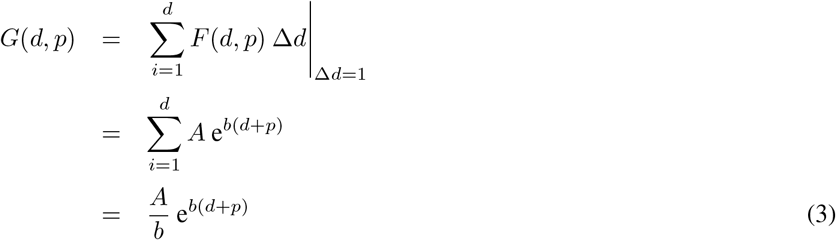

If *F* represents a cumulative figure, then its value at a particular time is:

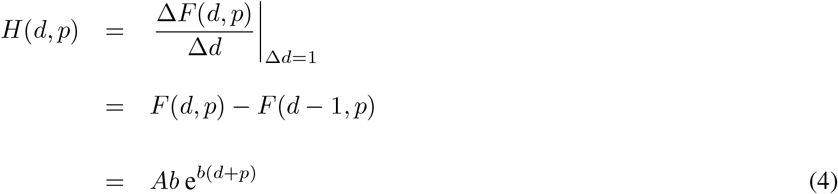

where Δ*d* = 1 day.

### D. Implementing the Model for Novel Coronavirus

#### 1) Modeling Cases for the World excluding China

To model the coronavirus outbreak in the world outside of China, a curve-fit is performed on the data collected.

The resulting equation for cumulative cases has the coefficients *b* = 0.201 and *A* = 1.9 wiht the understanding that these coefficients will change as the outbreak matures:

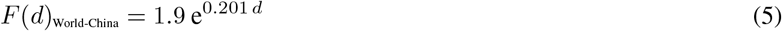

Figure 1 shows the correlation between the model and the case data for the world besides China.

**Fig. 1.**
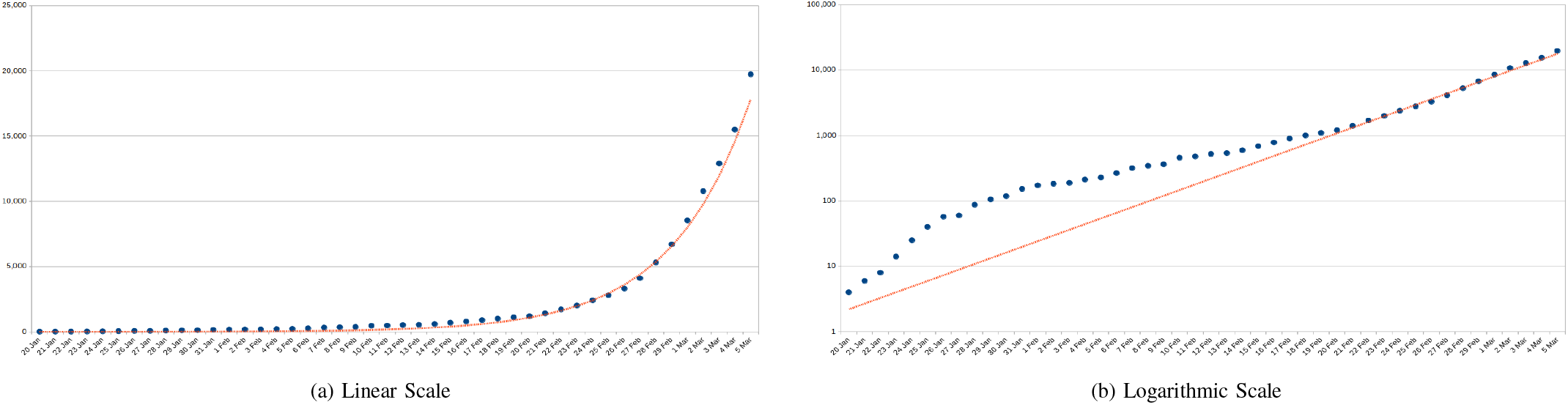
Cumulative number of confirmed coronavirus cases and the model for the world outside of China.

#### 2) Practical Assumptions to Model Novel Coronavirus

To simplify the model and frame the modeling space, the following assumptions are made:

1. Asymptomatic infections become contagious some time after the moment of infection from exposure.
2. The average person who has been infected will show symptoms after the average incubation period.
3. If testing could be 100% efficient, then every infected person will become a confirmed case statistic on average upon expiration of the average incubation period since being infected.
4. The model must react sensibly to external events.

Using these assumptions:

- The cumulative number of people infected on a given day are then estimated by shifting the curve into the past by the average incubation period.
- Inefficient testing percentages are accounted for by dividing the result by the testing percentage. That these two operations can be performed in any order.

### E. Modeling the Outbreak in the USA

#### 1) Initial Trend

Up until the end of February, the cumulative confirmed cases in the USA followed a function of the form:

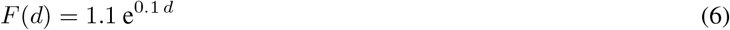

The rate of infection was initially significantly lower than the world average (outside of China). This can mainly be attributed to (i) imposing early flight restrictions on 31 January 2020 from heavily infected countries and (ii) testing had only been done case-by-case on people who showed symptoms and had entered the country from heavily infected areas. At this point, the outbreak was largely contained.

#### 2) Influence of Travel Restrictions

The imposition of early travel restrictions from China into the USA slowed down the virus by containing it mainly to individuals who had been to infected areas. The model indicates that delay was 26 days before it became “community-spread” in the USA. (Here, the term *community-spread* refers to a combination of person-to-person and surface-to-person transmission modes.)

The Figure 2 depicts what happened to the cumulative number of cases before and after the outbreak in the USA became community-spread. At that point, the trend line followed the same slope as the trend line for the world outside of China.

**Fig. 2.**
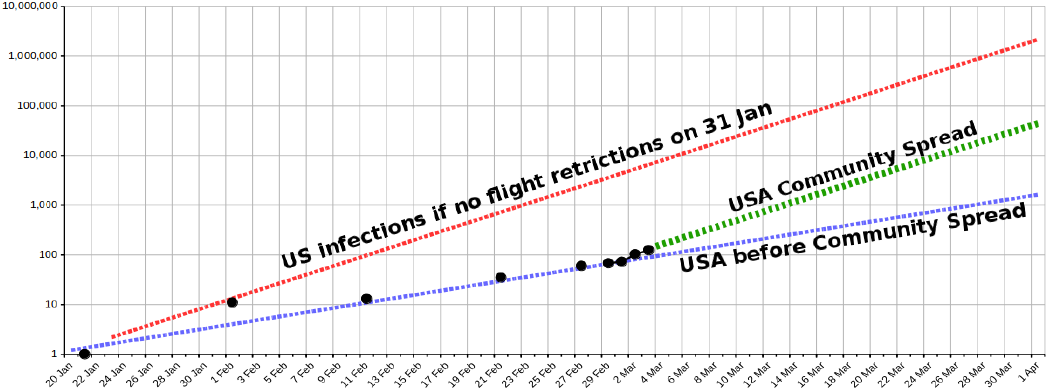
USA confirmed cases, the USA models before and after *community-spread* mode, and the model for the world outside of China.

#### 3) Influence of Testing Policies

During the first week of March 2020, the USA started expanding testing beyond their initial requirements. Testing was approved for people showing symptoms who did not have direct contact with people who had entered the country from infected areas. Production of testing kits in mass numbers began as well as distribution to the state laboratories. By this time, the virus had already left containment.

It is reasonable to assume that the number of cases being confirmed were lagging the actual number of people becoming ill due to inadequate testing. All the plots that follow are referenced to 29 February. As tests were being ramped up to catch up with the rate at which infections were becoming symptomatic and being reported, a surge in data was to be expected. Therefore, the model was *not* adjusted to follow the surge. The model having the slope equal to that for the world outside of China was still being used after 29 February even though the rate at which actual new cases were being confirmed in USA was higher. This ensured that the projected estimates of unreported infections weren’t artificially being inflated due to the lack of initial test data.

### F. Introducing Testing Efficiency

Testing efficiency, a percentage, can now be defined as follows:

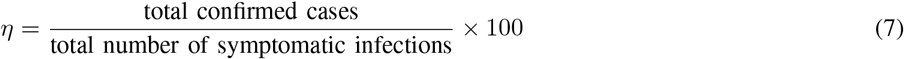

It represents the percentage people who have been tested positive out of all people who should have tested positive. It incorporates other parameters such as testing accuracy. Note that an incubation period has to expire for a person to become symptomatic and be tested positive. This will be treated in more detail shortly.

The fact of the matter is that without good data, there is no good way to estimate *η*. Without data it isn’t even possible to establish an appropriate range of values for *η*. It is therefore our approach to use *η* as a variable. The total number of symptomatic infections are:

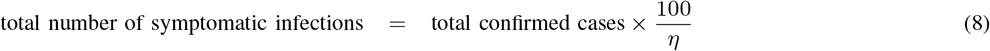

This means that a fraction *η* out of every 100 new cases were tested, and tested positive. (The accuracy of the test may be less than 100% and is also incorporated.) To understand this it might help to think about it from a different perspective. If out of all the people who became symptomatic on any specific day, a fraction *η* of them were tested and also tested positive, then the ratio of confirmed cases to the cumulative numbers for actual cases will remain *η*.

Using our model with *p* = 0, let *S* now represent the cumulative number of symptomatic infections in USA. Then:

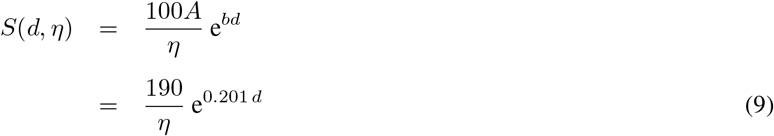

This is plotted in Figure 3(a), showing the total number of people who were infected and are showing symptoms to the point that they should have been tested.

**Fig. 3.**
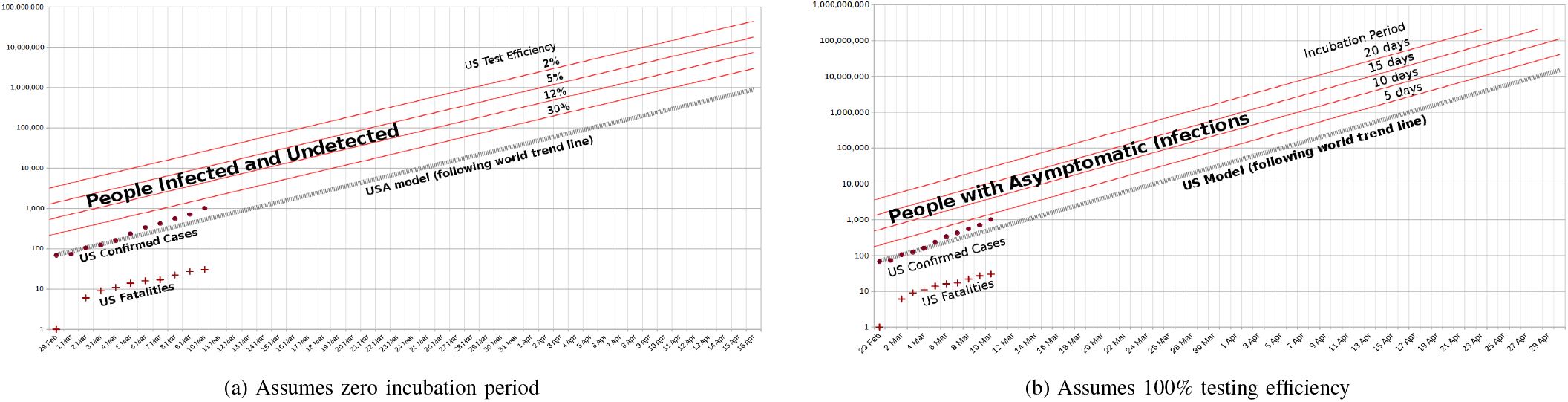
The total number of infected people in USA as function of US testing efficiency only and incubation period only.

#### 1) Introducing Incubation Period

Setting *η* aside for the moment, let’s now consider incubation period, a characteristic of the virus that we’ve defined as the average period (in days) between a person becoming infected to the onset of symptoms of illness.

To incorporate it into the model, let *I* represent the cumulative number of asymptomatic infections, and *p* the incubation period. Then:

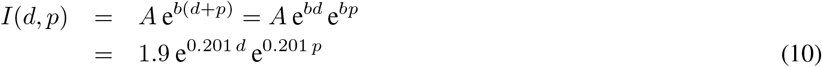

In a perfect world of testing, and if all cases operate at the averages, then every case added on a particular day will become a confirmed case after the incubation period. The curve for new cases confirmed would be precisely the same as the curve for the new infections, except for lagging it exctly by the average incubation period. The cumulative rates of new infections would then also lead the cumulative cases confirmed by the average incubation period. This is illustrated in Figure 3(b), showing the total number of asymptomatic infections for average incubation periods of 5, 10, 15, and 20 days.

### G. Incorporating both Incubation Periods and Testing Efficiencies

Combining the adjustment for the incubation period and the testing efficiency (in any order) into the model, the total number of infected individuals can be estimated. Let *P* now represent the cumulative number of infections. Then:

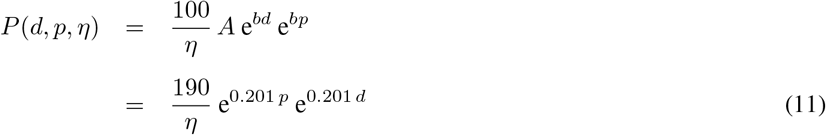

An incubation period of 4 days is assumed in Figure 4(a), which plots estimates of the total number of infected in the USA as a function of *η* = 2%, 5%, 12%, and 30%.

**Fig. 4.**
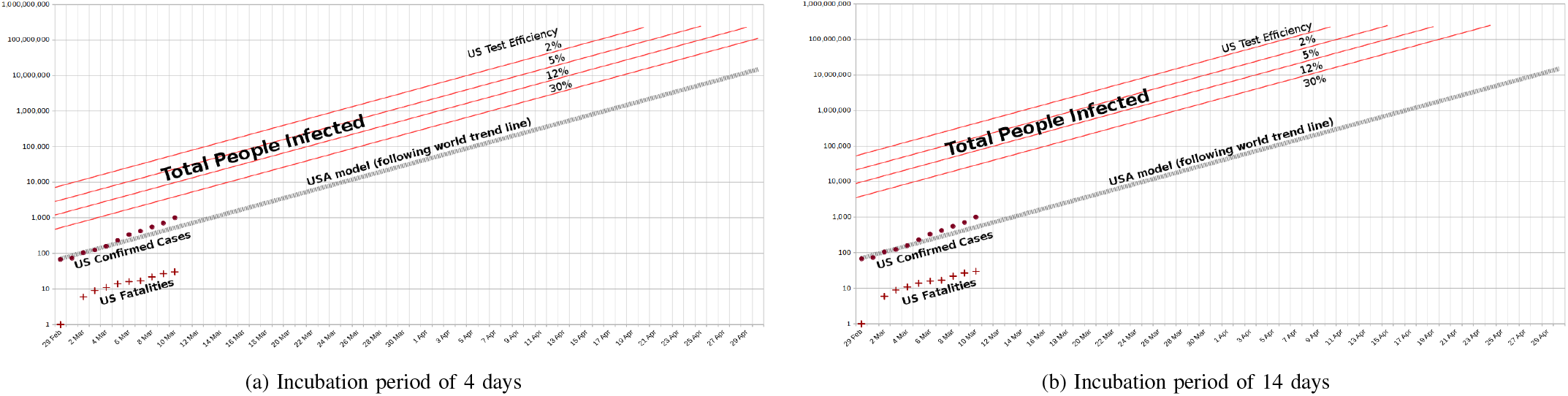
Total number of people infected in the USA as a function of testing efficiency and incubation period.

The way to read the graph, is by choosing the date-referenced testing efficiency, *η*, and starting with the corresponding red curve. This curve represents on the y-axis the cumulative number of people (on average) who are infected on the x-axis date. On average the same number of people became symptomatic (and testable) by *p* number of days later (*p* = the incubation period.) This can be plotted by moving the original red line to the right by *p* days. The line approximating the confirmed case data is then obtained by multiplying the last line by percentage *η*.

Figure 4(b) repeats it and assumes an incubation period of 14 days.

## Data Availability

All data is publicly available.

https://coronavirus.gov

